# Magnetic activated cell-sorting identifies a unique lung microbiome community associated with disease states

**DOI:** 10.1101/2021.06.02.21258247

**Authors:** Daniel G. Dunlap, Libing Yang, Shulin Qin, John Ries, Kelvin Li, Adam Fitch, Laurence Huang, Bryan J. McVerry, Barbara A. Methé, Alison Morris

## Abstract

**Rationale:** The advent of culture-independent, next-generation DNA sequencing has led to discovery of distinct lung bacterial communities. Studies of lung microbiome taxonomy often reveal only subtle differences between health and disease, but microbial host response may distinguish members of similar communities in different populations.

**Objectives:** Magnetic-activated cell sorting (MACS) has been applied to the gut microbiome to identify numbers and types of bacteria eliciting a humoral response. We adapted this technique to examine populations of immunoglobulin-bound bacteria and investigate the lung microbiota in HIV as a representative disease.

**Methods:** 42 people living with HIV (PLWH) and 22 HIV-uninfected individuals underwent bronchoalveolar lavage (BAL). We separated immunoglobulin G-bound bacteria using MACS and sequenced the 16S rRNA gene on the Illumina MiSeq platform. We analyzed sequences and quantified BAL cytokines and bacterial rRNA copy numbers.

**Measurements and Main Results:** Immunoglobulin G-bound bacteria were detectable in the healthy lung microbiota. Comparison of raw BAL by HIV status showed no significant taxonomic differences, but the immunoglobulin-bound lung microbiota differed by HIV status with greater abundance of *Pseudomonas* in PLWH. BAL cytokine levels were also higher in PLWH, which correlated with increased quantity of immunoglobulin G-bound bacteria.

**Conclusions:** We report a novel application of magnetic-activated cell sorting to identify immunoglobulin G-bound bacteria in the lung. This technique identified distinct bacterial communities which differed in composition from raw BAL, revealing differences in health and disease not detected by traditional analyses. Cytokine response was also associated with differential immunoglobulin binding of lung bacteria, suggesting functional importance of these communities.

## INTRODUCTION

Studying the lung microbiome is challenging given its low biomass, the propensity to detect contaminating bacteria using next-generation sequencing techniques, and the inability to determine functional impact of pulmonary microbes (1, 2). As a result, studies that compare the lung microbiome in different diseases such as COPD or HIV often have conflicting conclusions or find few major differences between bacterial communities (3).

Magnetic-activated cell sorting (MACS) can be used to separate and identify immunoglobulin-bound bacteria. Bacteria are stained with fluorophores specific for bacterial DNA and the Fc region of any immunoglobulin adhered to the bacterial cell wall. Magnetic labeling of stained bacteria allows IgG-bound and un-bound bacteria to be separated and analyzed. Recently, this technique has been applied to studies of the gut microbiome, and immunoglobulin (Ig)-bound bacteria have been found to modulate inflammatory bowel disease (4, 5). This technique has not been applied to the lung, but could serve to distinguish bacteria provoking an immune response versus those that are merely “innocent bystanders.”

We adapted the MACS technique from gut studies and applied it to the lung and bronchoalveolar lavage (BAL) in order to determine if IgG-bound bacteria differ from unsorted (raw) bacteria. We used human immunodeficiency virus (HIV) as a representative disease to evaluate differences in IgG-bound bacterial communities in the lungs of individuals with and without HIV infection. We performed quantitative polymerase chain reaction (qPCR) and next-generation sequencing on the 16S rRNA gene in IgG-bound and unsorted (raw) BAL samples. In addition, we quantified lung cytokine levels from raw BAL. We hypothesized that IgG-bound bacteria would be different from the overall bacterial communities and that people living with HIV (PLWH) would have increased recognition of and subsequent response to the lung microbiome via binding of IgG to resident bacteria.

## METHODS

### Study participants and sample collection

We recruited individuals with and without HIV infection through the University of Pittsburgh HIV research study at the Pittsburgh, Los Angeles and San Francisco sites. All participants provided written informed consent. The University of Pittsburgh, University of California, Los Angeles, and University of California, San Francisco institutional review boards approved this study. We collected BAL samples via bronchoscopy as previously described (6). Participants were instructed to avoid any food or beverages beginning at midnight before bronchoscopy and abstain from smoking at least 12 hours prior to the procedure. Additional detail is available in the online supplement.

### Magnetic-activated cell sorting

We used magnetic-activated cell-sorting to separate IgG-bound and –unbound bacterial in BAL by creating a magnetic label on IgG-bound bacteria then passing treated BAL samples through columns embedded within a super magnet. A detailed description of the assay is available in the online supplement.

### Flow cytometry and quantitative polymerase chain reaction

Immediately following each MACS assay, flow cytometry was performed. Quantitative PCR amplification was also performed to quantify bacterial rRNA copy number as previously described. Additional details available in the online supplement.

### Luminex panels and cytokine analysis

We used a commercially available Luminex assay (Bio-Plex Pro™ Human Cytokine 17-plex Assay, Bio-Rad Laboratories, Hercules CA) to quantify cytokines present within BAL, per manufacturer’s instructions. The cytokines used for analysis included interleukin (IL)-1β, IL-2, IL-6, IL-8, macrophage inflammatory protein (MIP-1B, also known as CCL4), interferon (IFN)-γ, and tumor necrosis factor (TNF)-α. No dilutions were performed.

### Sample processing and sequencing

We extracted bacterial DNA from both sorted and unsorted BAL samples using DNeasy PowerSoil Kits (Qiagen, Germantown MD) per manufacturer’s instructions, then amplified the V4 subunit of the bacterial 16S gene using polymerase chain reaction (PCR). We performed sequencing on the Illumina MiSeq platform (7) and de-multiplexed reads onboard the machine using standard Illumina software. We then performed post-sequencing quality control (including sequence filtering and trimming raw 16S sequence) using an in-house pipeline developed by the University of Pittsburgh Center for Medicine and the Microbiome, utilizing available software including fastx toolkit, cutadapt, and dust (8–10). Contamination controls were performed at every step of the process. Distribution-based contaminant filtering was applied to column controls to remove background signal from MACS columns and beads (Supplement 1). We generated operational taxonomic units (OTUs) and RDP classified sequences using an in-house mothur pipeline (11, 12).

### Quantifying immunoglobulin levels

Both serum and BAL immunoglobulin G levels were quantified. Serum samples were sent to the University of Pittsburgh Medical Center Clinical Laboratory (3460 Fifth Ave, Pittsburgh, PA), who quantified levels using commercially available ELISA assays. To quantify BAL IgG levels, we used a commercially available ELISA assay (Human IgG ELISA Kit, Abcam, Cambridge, MA) using manufacturer’s instructions, diluting BAL at a 1:500 ratio.

### Statistical analysis

First, we compared the IgG-bound and raw lung microbiota in HIV-uninfected individuals. Next, all participants were stratified by HIV status and use of anti-retroviral therapy (ART) in PLWH. Additional details available in the online supplement.

## RESULTS

Twenty-two HIV-uninfected participants and 42 PLWH were included. Age was similar between groups while gender, race, and smoking status differed (Table 1). Sixty-four IgG-bound, IgG-unbound, and raw samples were analyzed, though one raw sample had zero reads and was excluded from subsequent 16S analysis. IgG-unbound sorted aliquots largely resembled column contamination and were thus excluded from subsequent analysis.

**Table 1:**
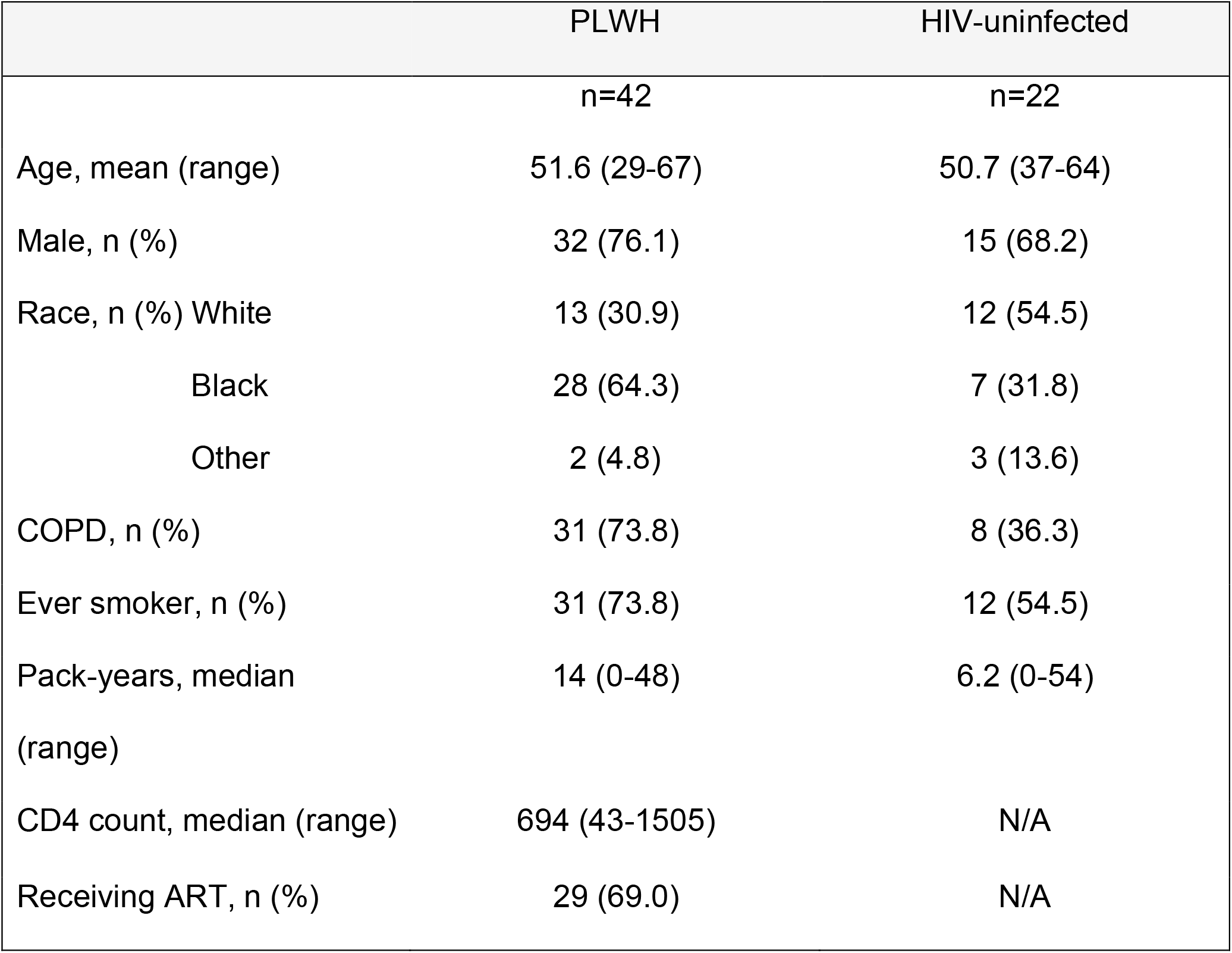
Participant Demographics by HIV Status (n=64)

### Comparison of IgG-bound lung microbiota to raw BAL microbiota in healthy, HIV-uninfected individuals

The relative abundance and overall community structure differed in IgG-bound and unsorted samples in HIV-uninfected individuals (PERMANOVA for Bray-Curtis distance, p<0.0001, R^2^=0.09; Figure 1A). While similar bacteria were seen in both raw and IgG-bound BAL samples, their relative abundance differed (Figure 1B). *Streptococcus, Prevotella*, and *Veillonella* were more abundant in raw BAL samples relative to IgG-bound BAL. These oral microbes were also seen in IgG-bound samples, but *Pseudomonas* was detected in greater relative abundance when compared to raw BAL samples.

**Figure 1:**
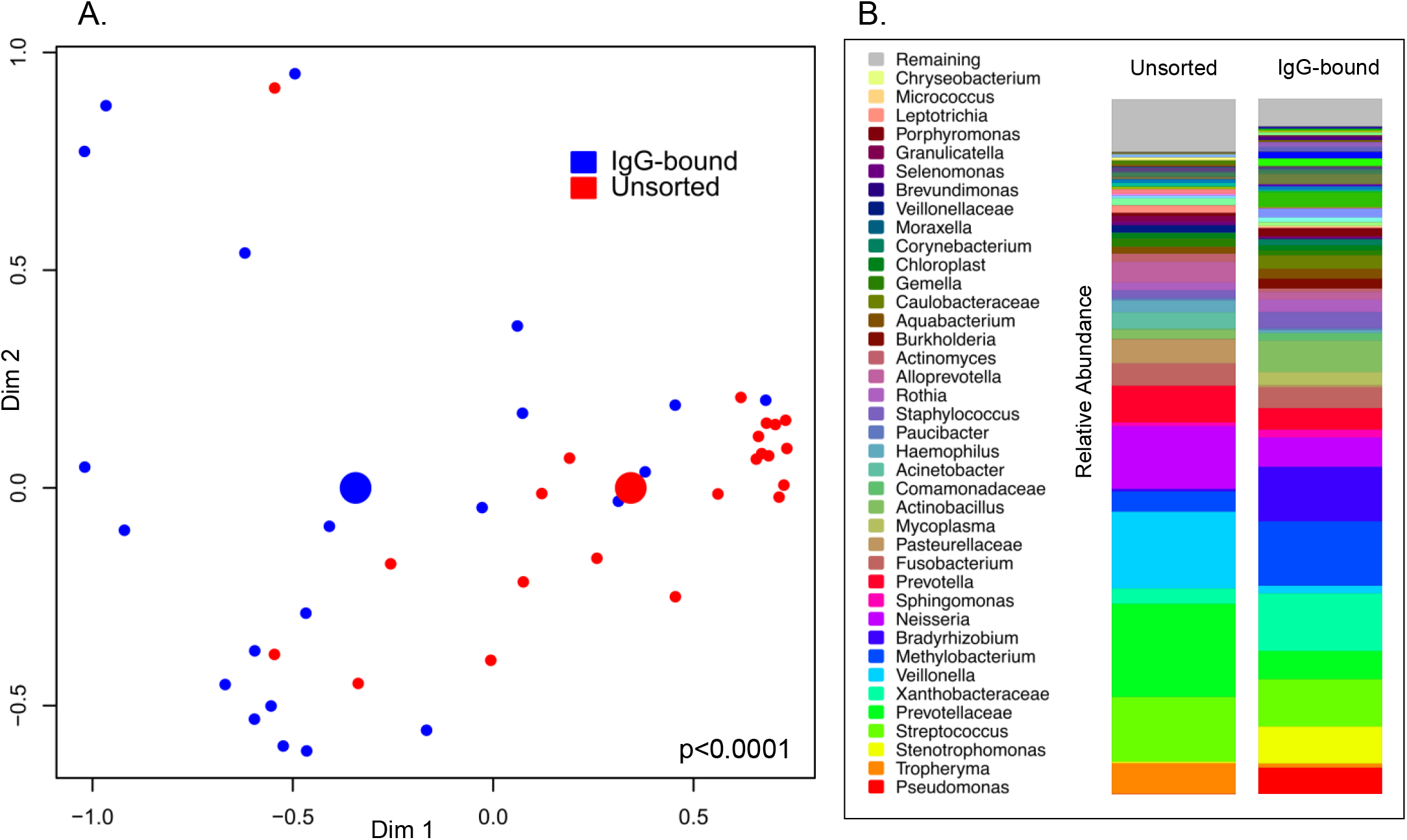
MACS identifies an IgG-bound lung microbiome that differs from raw BAL in healthy individuals. A non-metric multi-dimensional scaling plot (NMDS) (A) and stacked bar plots (B) detail the differences observed between the IgG-bound and raw lung microbiome in 22 HIV-uninfected individuals. **A)** We applied an NMDS plot using Bray-Curtis dissimilarity index to visualize sample clustering and performed multivariate analysis of variance (PERMANOVA) to compare beta diversity between IgG-bound and raw (unsorted) samples from 16S gene sequencing data. Points represent bacterial communities of individual samples, color-coded by IgG status (blue=IgG-bound, red=unsorted/raw). Larger colored centroids represent the mean group values. We found significant taxonomic differences between groups (p<0.0001, R^2^=0.09). **B)** Stacked bar plots depict the relative abundance of bacteria in unsorted and IgG-bound BAL samples from HIV-uninfected individuals. In raw BAL samples, common oral microbes such as *Streptococcus, Prevotella*, and *Veillonella* were most abundant. Relative abundance of bacteria differed in the IgG-bound fraction.

### Comparison of raw and IgG-bound BAL by HIV status

We then compared lung bacterial communities by HIV status and participant use of ART using flow cytometry data, 16S results, and qPCR. Based on flow cytometry, PLWH had more IgG-bound bacteria in the lungs than HIV-uninfected individuals (p=0.0008, Figure 2A). Further, PLWH not on ART tended to have a higher quantity of IgG-bound bacteria than those on ART. PLWH not on ART had greater IgG-bound bacteria in BAL than persons without HIV (p<0.0001, Figure 2B). Utilizing qPCR to quantify the number of bacterial 16S rRNA copies in IgG-bound BAL samples, PLWH tended to have higher IgG-bound rRNA copy numbers than HIV-uninfected individuals (p=0.06, Figure 2C). We also quantified serum and BAL IgG concentration. There was no difference in IgG concentration in PLWH compared to HIV un-infected individuals, but PLWH had high serum IgG level (p=0.03, Supplement 2).

**Figure 2:**
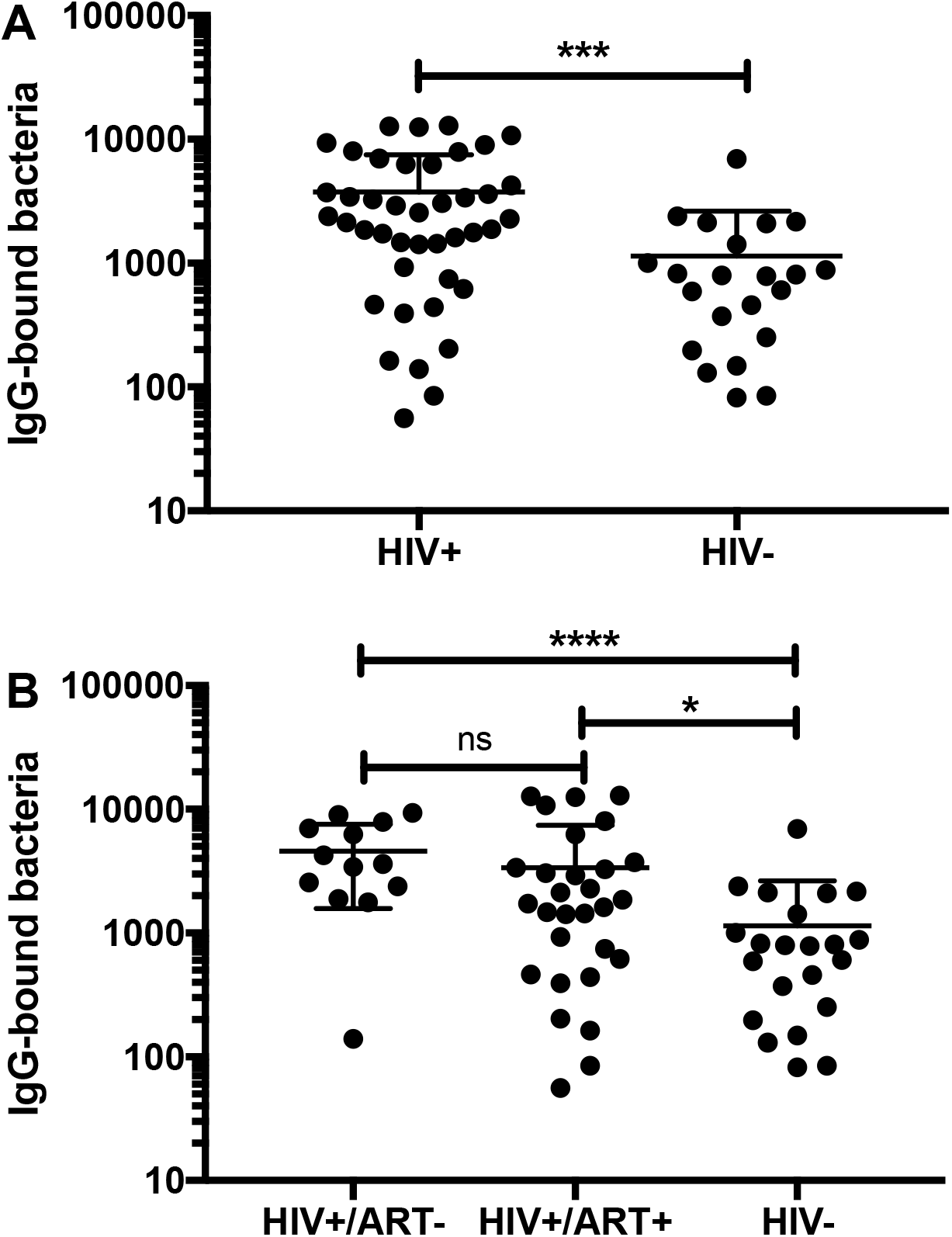
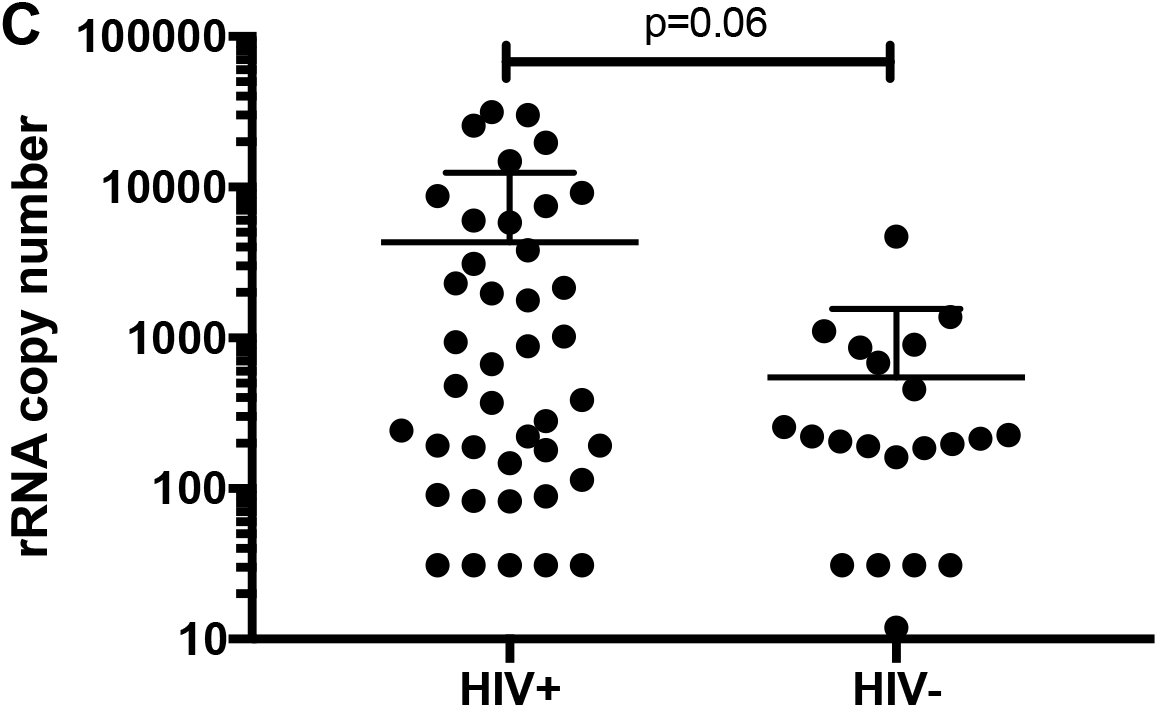
PLWH had greater number of IgG-bound bacteria in the lungs than HIV un-infected individuals. Study participants were grouped by HIV status and then by use of anti-retroviral therapy (ART). **A)** Individuals were grouped by HIV status and groups compared using non-parametric t-testing (Mann U Whitney). PLWH had significantly more IgG-bound bacteria than HIV-uninfected individuals (p=0.0008). **B)** PLWH were then sub-divided by use of ART and compared with HIV-uninfected individuals. The three groups were compared using non-parametric t-tests (Mann U Whitney). PLWH not receiving ART had the highest abundance of IgG-bound bacteria by flow cytometry, when compared to HIV-uninfected individuals (p<0.0001) and PLWH taking ART (p=0.06). PLWH on ART also had greater abundance of IgG-bound bacteria (p=0.017). **C)** Quantitative PCR was used to quantify rRNA copy number in IgG-bound BAL samples. PLWH tended to have higher rRNA copy number (p=0.06).

Next, we compared BAL microbial composition in PLWH and HIV-uninfected individuals. Similar to previous studies (3), we found no significant differences in the lung microbiota when comparing PLWH to HIV-uninfected individuals (Figure 3A). However, when we performed next-generation sequencing on IgG-bound samples, bacterial communities stratified by HIV status (PERMANOVA for Bray-Curtis distance, p=0.008, R^2^=0.03; Figure 3A). In PLWH, *Tropheryma, Veillonella*, and *Prevotella* were the most abundant bacteria in raw BAL samples. In contrast, *Pseudomonas* was by far the most abundant IgG-bound bacteria in PLWH. In both HIV-uninfected individuals and PLWH, *Pseudomonas* was relatively more abundant in IgG-bound BAL samples when compared to raw but was significantly more abundant in PLWH (Figure 3B).

**Figure 3:**
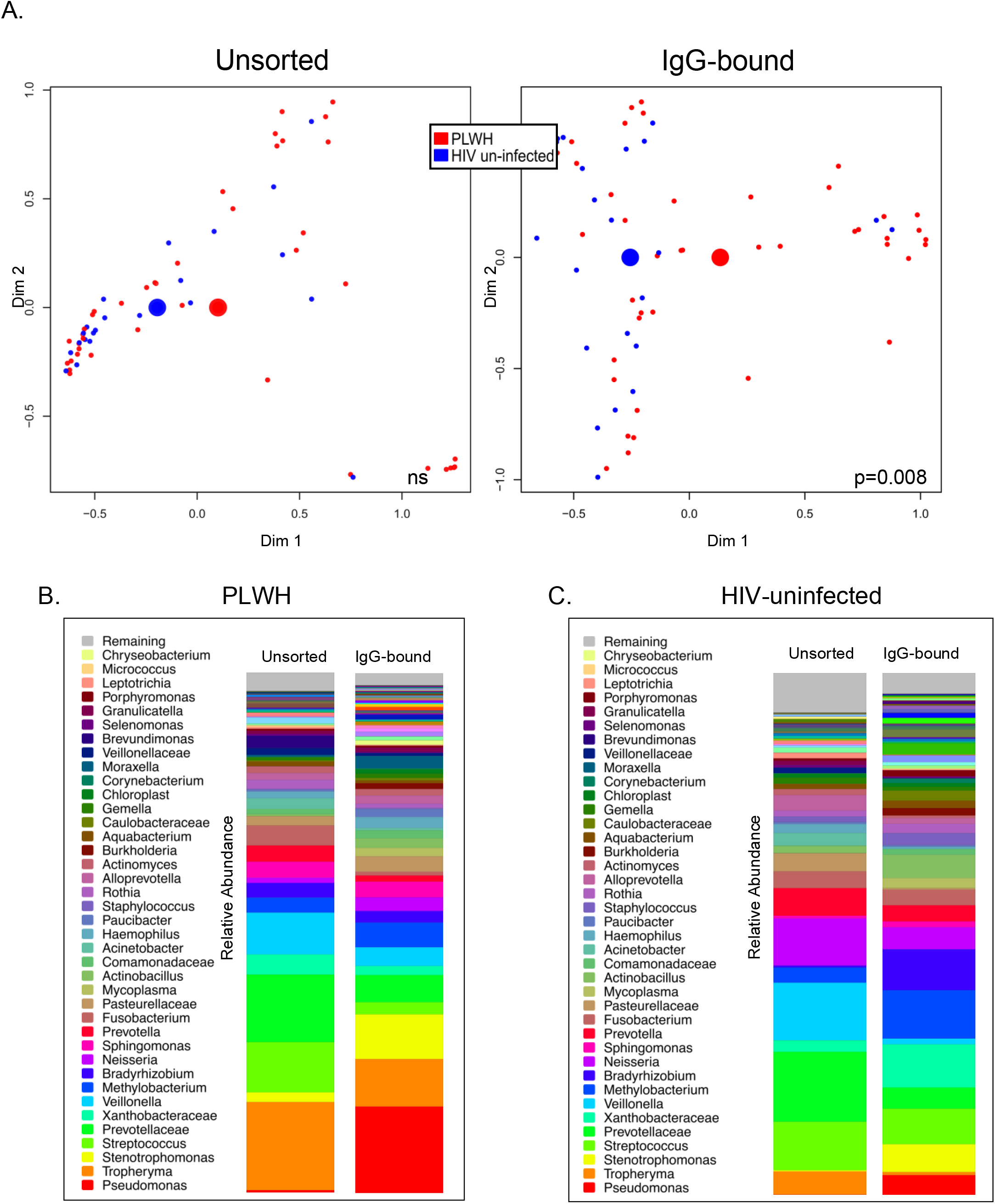
Bacterial taxa stratify by HIV status in IgG-bound BAL. **A)** Unsorted (left) and IgG-bound (right) NMDS plots are displayed side by side to compare the differences in beta diversity observed when individuals were grouped by HIV status. We used Bray-Curtis dissimilarity index to visualize sample clustering and performed multivariate analysis of variance (PERMANOVA) to compare bacterial communities in PLWH and HIV-uninfected individuals in both unsorted (raw) and IgG-bound BAL samples. Smaller dots signify the microbial community of individual samples, while the larger centroids represent the average of each community (red=PLWH, blue=HIV un-infected). There were no taxonomic differences between PLWH and HIV un-infected individuals in unsorted samples; however, bacterial communities stratified by HIV status in IgG-bound BAL samples (p=0.008). **B)** Stacked bar plots depict the relative abundance of bacteria in unsorted and IgG-bound BAL samples in PLWH. *Pseudomonas* was seen in greater abundance in IgG-bound samples when compared to unsorted samples. **C)** Stacked bar plots depict the relative abundance of bacteria in unsorted and IgG-bound BAL samples in HIV-uninfected individuals.

### Relationship between lung cytokine levels and IgG-bound bacteria in HIV

To further explore an association between host recognition of the lung microbiome and inflammatory response, we quantified BAL cytokine levels in the PLWH and linked these to levels of IgG-bound bacteria. We found that PLWH had significantly higher levels of BAL cytokines including IL-8, IFN-γ, MCP-1 and TNF-α as previously described (13). IL-6 and IL-1β levels also tended to be higher in PLWH (Supplement 3). In PLWH, higher levels of IgG-bound bacteria were associated elevated BAL cytokine levels including IL-8 and IL-1β by qPCR (p=0.003, R^2^=0.2 and p=0.012, R^2^=0.15, respectively) and flow cytometry (p=0.008, R^2^=0.16 and p<0.001, R^2^=0.32, respectively; Figure 4). There was no correlation between IL-8 and IL-1β cytokine levels with IgG-bound bacteria in HIV-uninfected individuals.

**Figure 4:**
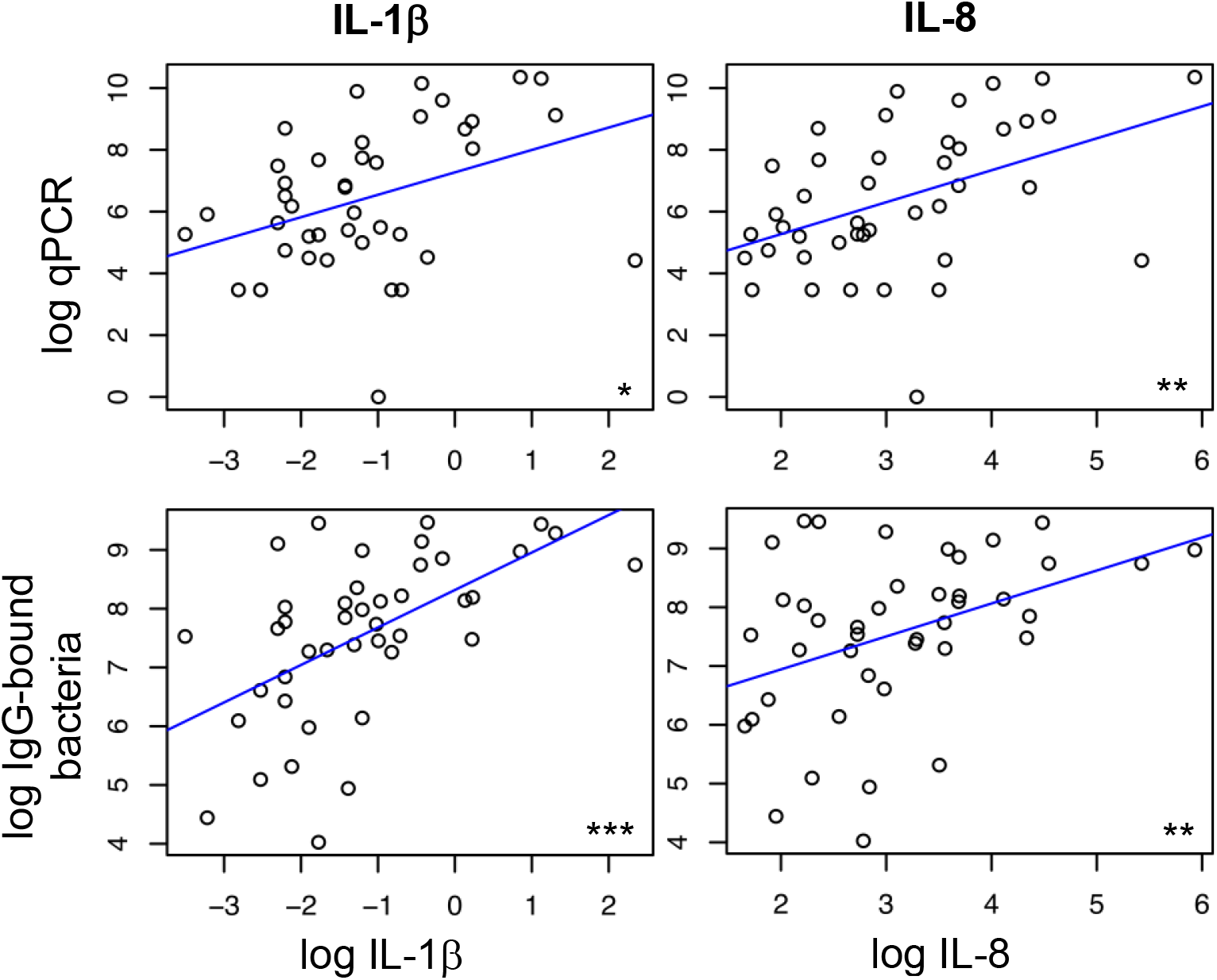
Increased IgG-bound bacteria in the lungs of PLWH correlate with BAL cytokine levels. We plotted IgG-bound bacteria as determined by qPCR (top) and flow cytometry (bottom) against BAL cytokine levels in PLWH. Individual dots represent the intercept between log-transformed cytokine level (x-axis) and log-transformed IgG bound bacteria quantity (y-axis). A linear regression model was applied to assist with visualization of the trend and quantify the degree of association. IL-8 and IL-1β correlated with increasing quantity of IgG-bound bacteria by qPCR (p=0.003, R^2^=0.2 and p=0.012, R^2^=0.15, respectively) and flow cytometry (p=0.008, R^2^=0.16 and p<0.001, R^2^=0.32, respectively).

## DISCUSSION

Using a novel application of magnetic-activated cell sorting to isolate IgG-bound bacteria and stufy the lung, we identified a distinct immunoglobulin-bound lung microbiota. Microbes bound by IgG were present in different relative abundance, resulting in markedly different community structure. Despite relatively high abundance of *Tropheryma* and oral anaerobes in unsorted samples, *Pseudomonas* was the most abundant bacteria recognized and bound by IgG. Using this technique, we detected differences in the lung microbiota in HIV infection that were not apparent when performing typical 16S rRNA gene sequencing of whole BAL. Further, IgG-bound bacteria in HIV were associated with increased inflammation within the lungs. Application of this technique may expand investigation of the lung microbiome in health and disease.

Initial work with MACS has been applied to study bacteria in the gut and gastrointestinal disorders. For example, using a murine model, researchers demonstrated that IgA-bound bacteria resulted in increased susceptibility to colitis in germ-free mice (4). In a subsequent human study, IgG binding identified bacteria implicated in inflammatory bowel disease pathogenesis in pediatric patients (5).

In the first investigation applying the MACS technique to the lung, we were able to isolate and quantify IgG-bound bacteria and reliably detect a different bacterial community structure than revealed by 16S rRNA gene sequencing of whole BAL. Although the typical oral bacteria identified in the lung microbiome were seen in both IgG-bound and raw BAL, there was relatively greater IgG-binding of *Pseudomonas*. Functional significance of immunoglobulin-bound bacteria is not known, but suggests differences in host response to similar bacteria and may contribute to previously defined “pneumotypes” (14).

We investigated HIV as a representative condition for applying this technique to the study of pulmonary disease, and we were able to detect clear differences that were not seen in analyses of whole BAL. Prior investigation of the lung microbiome in HIV infection has failed to find significant taxonomic differences in individuals with normal CD4 counts on appropriate antiretroviral therapy (1, 3); a somewhat unexpected finding given the differences in lung immune responses and pulmonary diseases in this population (15–19). PLWH had greater abundance of IgG-bound respiratory pathogens such as *Pseudomonas* and *Stenotrophomonas*. Elevated levels of multiple cytokines, including IL-6 and IL-1β, were correlated with higher numbers of IgG-bound bacteria. This correlation suggests that host recognition of the lung microbiota may stimulate chronic lung inflammation. It is possible that HIV-infected individuals have increased recognition of the lung microbiota via binding of IgG to resident bacteria, leading to an increased inflammatory response and ultimately contributing to chronic pulmonary disease seen in this population (15, 17, 19). Future studies will be useful to examine this link.

Interestingly, *Tropheryma* was identified in raw BAL samples in both PLWH and healthy controls, but it was not as abundant in the IgG-bound communities. This bacteria has been consistently detected in the lung in both healthy populations and in PLWH (20). It has been associated with smoking and its prevalence decreases with initiation of ART. Despite its presence in the lung, it has not been associated with pulmonary inflammation or lung function (21). The discordance between its high abundance in raw samples and relatively lower abundance in IgG-bound samples could suggest that the absence of an inflammatory response to *Tropheryma* in the lungs may be due to a lack of active host recognition and response (20, 21).

It is plausible that immunoglobulin-bound bacteria play a role in lung disease. Though traditionally considered protective, immunoglobulin binding of bacteria can lead to inflammation through opsonization (22) and antibody-dependent cell-mediated cytotoxicity (23). B cells have been identified in greater abundance from the lungs of individuals with severe COPD (24) and have been implicated in emphysema pathogenesis (25, 26). Host recognition of the lung microbiome corresponds to an inflammatory response (14). Our results suggest that one possible mechanism of host recognition and response to the microbiome is through the adaptive immune system. Further, the presence of IgG-bound bacteria in the lungs given the relative absence of IgG in the mouth could suggest an alternative mode of bacterial migration into the lungs or recognition of unbound bacteria in the lungs.

Our study has several limitations. Given the low biomass within the lungs, any degree of experimental contamination can significantly skew results. We attempted to combat the impact of contamination by filtering all media, running UV light in the biosafety cabinet at the beginning and conclusion of each assay, and performing controls at each step of our experiment. We ran MACS controls, where 1% buffer was stained and run through the columns and subsequently sequenced. Using the sequencing data from the columns, we were able to subtract the 16S signal attributable to column contamination, reducing the amount of background noise. Additionally, controls were performed during each step of the PCR process. We examined only IgG, but IgM or IgA binding might identify other bacterial communities of importance. We chose IgG as our initial analysis based on preliminary fluorescence-activated cell sorting data demonstrating the strongest signal (data not shown). Other variables such as smoking or prior lung infections could also impact results.

In conclusion, we report the first study of the immunoglobulin-bound lung microbiome. Using this technique, we identified distinct bacterial communities in the healthy lung and in HIV-infected individuals. IgG-bound bacteria showed greater differences by HIV status than seen in traditional analyses of BAL, and the relationship to inflammation suggests a potential role in disease. Though we chose to study immunoglobulin binding in HIV, adaptive immunity has been implicated in myriad chronic pulmonary diseases (24, 27, 28), thus study of host recognition and response to the lung microbiome can be applied to investigation of other lung diseases, and modulation of immunoglobulin-binding of the lung and oral microbiome could be a therapeutic target.

## Data Availability

All sequencing data is available at https://www.ncbi.nlm.nih.gov/sra. Project name: PRJNA720126.

https://www.ncbi.nlm.nih.gov/sra/?term=PRJNA720126

