## Supplemental File for "Magnetic activated cell-sorting identifies a unique lung microbiome community associated with disease states"

### Methods – Additional Detail

#### Study Participants

Individuals had no acute respiratory symptoms at the time of enrollment and were specifically without fevers, upper respiratory infection, or lung infection. Exclusion criteria also included those who had received systemic corticosteroids within the previous 6 months and individuals receiving antibiotic therapy within 3 months of participation. As individuals had previously participated in a variety of studies of pulmonary disease in HIV (1), we included those with sufficient volumes of BAL for evaluation. All participants provided written informed consent. The University of Pittsburgh, University of California, Los Angeles, and University of California, San Francisco institutional review boards approved this study.

**Sample Collection:** All subjects gargled an antiseptic just prior to instrumentation in an attempt to limit bacteria in the oropharynx. Before bronchoscopy, 10 to 50 mL 0.9% normal saline was flushed through the bronchoscope and used as a negative control. Under moderate sedation, the bronchoscope was passed through the vocal cords and quickly placed into the wedge position in the right middle lobe. We collected BAL using sterile 0.9% normal saline to a maximal instillation volume of 200 mL. BAL specimens were immediately fractioned into 1 and 5 mL aliquots and stored at -80ºC.

#### Flow Cytometry

Immediately following each MACS assay, flow cytometry was performed. Four hundred-fifty μL from IgG-bound and IgG-unbound aliquots were stored at 4ºC for no more than

2 hours. After performing stain compensation on the STI Fortessa (STI Electronics, Madison, AL), all samples were run at high throughput speed for 4 minutes. Plotting side scatter (SCC-A) against fluorescein isothiocyanate (FITC, emission 488nm) was used to gate the appropriate population of interest. A second plot of FITC vs phycoerythrin (PE, emission 578nm) was performed. Gated reads with both a positive FITC and PE signal ($<10^3$) were quantified, representing the relative amount of IgG-bound bacteria in a given sample. Negative column controls, which contained only processed, stained buffer were performed during sample runs as a quality control and used for subsequent 16S control.

**Magnetic-activated Cell Sorting**

One mL BAL and 1 mL buffer (1% BSA in PBS) were filtered into 2 cc Eppendorf tubes, and bacteria were isolated via centrifugation (8000 RPM x5 minutes). We removed a small amount of bacteria suspended in buffer to be used in compensation controls for flow cytometry at the conclusion of each sort (described below). Remaining bacterial pellets were suspended in a solution containing the bacterial DNA stain, SytoBC (Thermo Fisher Scientific, Waltham, MA) and the immunoglobulin stain, IgG-PE (eBioscience, Thermo Fisher Scientific, Waltham, MA). We incubated stained bacteria with anti-PE microbeads (Miltenyi Biotec, USA), then used centrifugation to purify magnetically-labeled, stained bacteria prior to magnetic-activated cell sorting (MACS).

After washing MACS columns (Miltenyi Biotec) with buffer, we poured individual bacterial samples into corresponding MACS columns that were embedded within a super magnet (OctoMACS Separator, Miltenyi Biotec). Fifteen mL conicals (IgG-unbound) were stationed beneath each column. Once the entire suspended bacterial sample passed through, each column was removed from the OctoMACS Separator, placed within a different IgG-bound conical, and flushed with 1 mL buffer. The buffer was vigorously plunged through the column, per manufacturer's instructions, removing the magnetically-labeled material that had been held in suspension during the assay. Columns were then removed from their respective IgG+ conicals and placed atop a vacuum manifold (Promega, Madison WI) where they were washed with 70% EtOH followed by buffer. Columns were replaced to their prior locations within OctoMACS Separator and the contents within the IgG- conicals were poured into the same columns now directly above. An additional washing step followed, with another 1 mL buffer plunged through the column into the same IgG+ conical, bringing the total volume within each conical (both IgG-bound and IgG-unbound) to 2 mL, 450 μL of which was separated and used for flow cytometry analysis.

**Quantitative PCR**

Quantitative PCR amplification was performed in a total volume of 20μL amplification reaction consisting of 2μL of 10XPCR buffer, 3.5 mmol/L MgCl2, 0.2 mmol/L deoxynucleoside triphosphate, 0.5 μmol/L forward and reverse primers, 0.225 μmol/L probe, 0.75 U of Platinum Taq polymerase (Invitrogen), and 2μL of each DNA. The forward and reverse primers were used to amplify DNA templates encoding 16S rRNA and both primer and probe sequences were identical to those previously described (2). A standard curve was created from serial dilutions of plasmid DNA containing known copy numbers of the template. The assays were performed on the LightCyler System

(Roche) using the following PCR conditions: 95°C for 5 min, followed by 50 cycles at 95°C for 15 s and at 60°C for 1 min.

**Statistical Analysis**

We applied a non-metric multidimensional scaling plot (NMDS) using the Bray-Curtis dissimilarity index to visualize sample clustering and performed multivariate analysis of variance (PERMANOVA) to compare the microbiota by HIV status, IgG status, ART status, and BAL cytokine level with the *R* vegan package. We performed paired testing of BAL samples between IgG-bound and unsorted samples using Wilcoxon paired p-values to detect differences in abundance of individual bacterial genre. Non-parametric t-testing (Mann U Whitney) was used to compare groups by HIV status and use of ART. Cytokine levels were log-transformed and plotted against log-transformed IgG-bound bacteria and qPCR data, and a linear regression model was used to evaluate correlation between the two variables.

**Supplement 1 - Distribution-based contaminant filtering**

The goal of computational contaminant filtering is to subtract the pattern of taxa found in the controls from the experimental samples. This represents a compromise between liberally removing all taxa found in the controls (increasing the likelihood of eventual false negative associations) versus performing no contaminant filtering whatsoever (increasing the likelihood of eventual false positive associations). The compromise involves estimating the proportion of each taxa that should be deemed contaminant and then removing them from the experimental measurements. The proportion to remove follows this mixture model:

**(1-p)*actExp + p*actCont = obsExp**

Each observed experimental (obsExp) sample is allowed to have its own proportion, p, of actual contaminant (actCont) and the remaining proportion (1-p) of actual experimental sample. Since we cannot measure the actual contaminant, we use the observed controls (obsCont) as an approximation. Each sample is a multivariate measurement of taxa abundance, and should be considered compositional. Thus, the mixture model assumes that the relative proportion of each taxa, introduced by the contaminant is fixed. By fitting this objective function, the parameter p can be estimated for each sample by minimizing this objective function:

**Sum(p*obsCont[i]-(1-p)*obsExp[i])^2**

obsCont[i] and obsExp[i] represent the relative abundance of the ith taxon among the observed control and observed experimental sample, respectively. This sum of squared difference objective function is minimized when the scaled shape of the observed control's distribution of taxa overlaps the shape of the observed experimental sample's distribution. Since there is an expectation that the observed experimental (obsExp) sample will contain taxa not found in the observed control (obsCont), these taxa are excluded from the objective function. This ensures that there is no mismatch penalty for the observed experimental sample containing taxa not found in the observed control. The same proportion p is applied to each taxa for a given sample. If an observed experimental sample is purely contaminant, then the objective function will minimize to

0, because the distribution of taxa in the observed experimental sample will look identical to the observed control sample.

After the contaminant proportion p is estimated, the proportion of the observed control is subtracted from the observed experimental. If the subtraction yields a negative value for a particular taxa, then its new abundance is set to 0. The resultant distribution of taxa is then normalized so that the sum across the relative abundance of all taxa equals 1. The calculated proportion is also used to scale down the read depth, so that the actual number of filtered reads per sample can be estimated.

When a control can be paired with an experimental sample, then the algorithm can be applied between the two members of the pair. However, in this experiment, where a smaller batch of controls was measured for a larger set of experimental samples, the average of controls was used as the observed control since the variation across controls was low and significantly different than the experimental samples.

**Supplement 2 – IgG quantification in blood and BAL**

We measured BAL and serum IgG levels and compared levels between PLWH and HIV uninfected individuals. There was no significant difference in BAL concentration between individuals with and without HIV infections (A, p=0.07). PLWH had higher serum IgG concentration (B, p=0.03).

A.

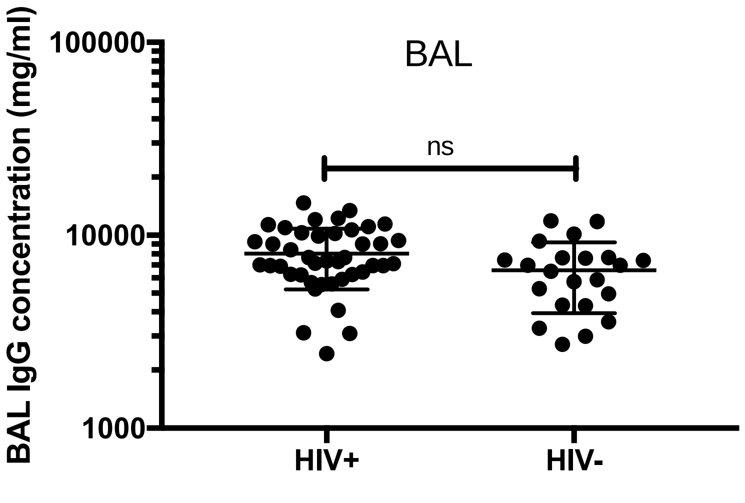
BAL
ns
100000
10000
1000
BAL IgG concentration (mg/ml)
HIV+
HIV-

B.

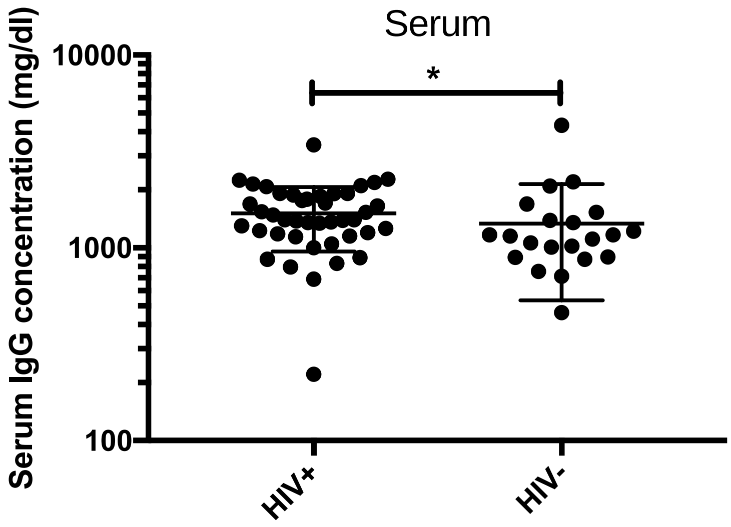
Serum
*
10000
1000
100
Serum IgG concentration (mg/dl)
HIV+
HIV-

**Supplement 3 – BAL cytokine levels**

We measured BAL cytokines and compared concentration between PLWH and HIV uninfected individuals. Non-parametric t-testing (Mann U Whitney) was used to compare groups. Dots represent BAL cytokine concentration (pg/ml) in individual samples. PLWH had higher levels of several cytokines implicated in COPD pathogenesis, including TNF-$\alpha$ (p=0.03), IL-8 (p=0.03), IFN-$\gamma$ (p=0.001), and MCP-1 (p=0.026). IL-6 and IL-1$\beta$ levels tended to be higher in PLWH, though this was not statistically significant (p=0.07 and 0.052, respectively).

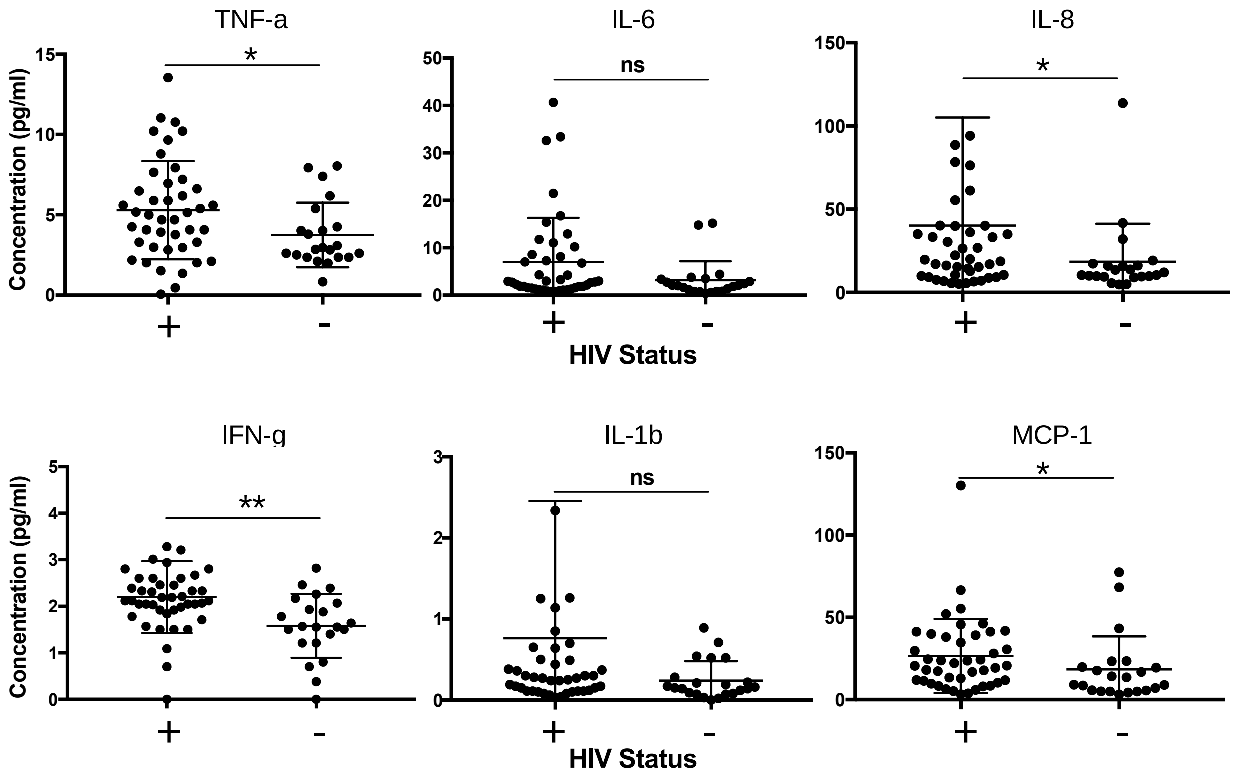
